# A Return-on-Investment Analysis of a Community-Based Diabetes Self-Management Program In New York City

**DOI:** 10.64898/2026.04.22.26351481

**Authors:** Jason Goldwater, Yael Harris, Sonali K. Das, Maria A. Fernandez Galvis, Duncan Maru, William B. Jordan, Crystal Sacaridiz, Chris Norwood, Sara Soonsik Kim, Kyle W. Neustrom

## Abstract

**OBJECTIVE:** To evaluate the return on investment (ROI) of a community-based Diabetes Self-Management Program (DSMP) enhanced with health-related social needs (HRSN) screening and referrals, implemented by the New York City (NYC) Department of Health and Mental Hygiene with three community-based organizations in highly-impacted, under-resourced neighborhoods.

**RESEARCH DESIGN AND METHODS:** A retrospective cost-benefit analysis from a public-sector payer perspective was conducted among 171 adults with type 2 diabetes who completed a six-week, peer-led DSMP delivered by community health workers (CHWs) in English, Spanish, and Korean during 2018–2019. A time-driven, activity-based costing model captured direct implementation costs, CHW workforce turnover, and administrative overhead. Monetized benefits included avoided diabetes-related complications, reductions in self-reported emergency department (ED) visits and hospitalizations, and quality-adjusted life year (QALY) gains from improved medication adherence. Univariate sensitivity analyses tested robustness under conservative assumptions.

**RESULTS:** Total program costs were $179,224; monetized benefits totaled $1,824,213, yielding a net benefit of $1,644,989 and an ROI of 918%—approximately $10 returned per $1 invested. Excluding QALY gains, ROI remained 551%. Self-reported ED visits declined from 149 to 82 and hospitalizations from 93 to 24 in the six months following intervention. Over 80% of participants reported housing instability; 72% were Medicaid-covered and 16% uninsured. Sensitivity analyses confirmed a positive ROI under all conservative scenarios.

**CONCLUSIONS:** A CHW-led, community-based DSMP integrated with HRSN screening and referrals delivered substantial economic and public health value among adults facing housing instability and structural barriers to care. Findings support inclusion of DSMP as a covered benefit in Medicaid managed care, value-based payment arrangements, and housing access initiatives to advance equitable diabetes outcomes.

## Introduction

As of 2022, over 37 million Americans were living with diabetes, with estimated national costs exceeding $327 billion annually, including $237 billion in direct medical expenses and $90 billion in lost productivity due to complications.^1^ The burden of diabetes is disproportionately concentrated among Black, Latino, immigrant, and lower household income populations^.2^

In New York City (NYC), diabetes is the third leading cause of death, with the highest prevalence in the Bronx (15%).^3^ Often related to historic segregation and its persistent effects in current day in the same communities,^4^ higher rates of health-related social needs (HRSNs), including housing instability, food insecurity, low health literacy, and limited access to primary care, contribute to the disproportionate burden of diabetes and its complications.^5^

The NYC Department of Health and Mental Hygiene (Health Department) partnered with three community-based organizations (CBOs), Health People Inc., Korean Community Services of Metropolitan New York Inc., and The Mexican Coalition for the Empowerment of Youth and Families, on a 2018-2019 pilot program funded by OneCity Health, under New York State’s Delivery System Reform Incentive Payment (DSRIP) program.^6^ To address the social and behavioral drivers of health disparities, they augmented the Diabetes Self-Management Program (DSMP), with social needs screening and referrals or people, a majority of whom face unstable housing. The DSMP is an evidence-based program associated with improved health outcomes for people with diabetes, including decreased all-cause emergency department visits, improved A1C (a measure of average blood glucose levels over 3 months), and medication adherence. The team incorporated a social needs screening component into this workflow to improve care coordination and address HRSNs among adults living with type 2 diabetes in highly-impacted, disinvested neighborhoods.

The pilot trained community health workers (CHWs) to deliver weekly 2.5-hour workshops over six weeks, supported by social needs screening and referrals to community services via the NowPow (now UniteUs) digital platform, with technical assistance provided to clinical and community-based service-delivery partners. The screening tool was constructed from previously validated items, filtered by demographics, and automatically generated referral recommendations. A mixed-methods evaluation of the program revealed meaningful improvements in participants’ diabetes knowledge, health literacy, and self-efficacy. The program achieved significant reductions in self-reported acute healthcare utilization: among the 171 participants, emergency department visits and hospitalizations decreased from 149 to 82 and from 93 to 24, respectively, during the 6 months following the intervention, compared with the 6 months preceding it.

This study presents a comprehensive return-on-investment (ROI) analysis of the DSMP conducted by the Health Department during 2018–2019. The analysis incorporates direct implementation costs, CHW workforce development, administrative overhead, and a range of monetizable benefits, including avoided diabetes-related complications, reduced emergency department and inpatient utilization, and quality-adjusted life years (QALYs) attributable to improved medication adherence. Additionally, enrollment in health insurance enhances diabetes self-management by reducing financial barriers to medications, glucose monitoring supplies, and primary care visits, yielding average annual savings of approximately $800 per person while reducing the risk of costly complications and uncompensated care.^7^ Using a participant-level costing approach, the study scales outcomes across 171 participants who completed the entire six week program to assess cost-effectiveness from a public health payer perspective. By quantifying the financial return of a community-based, equity-driven intervention, this analysis contributes to the growing literature on value-based prevention models. It highlights the importance of sustained investment in CHW-led chronic disease programs that address social determinants of health when serving highly impacted, under-resourced populations.

## Methods

This study employed a retrospective cost-benefit analysis framework to assess the ROI of the DSMP, coupled with social needs screening and referral, implemented by the Health Department and partners between 2018 and 2019. The evaluation followed established economic evaluation principles and adopted a public-sector payer perspective, quantifying both implementation costs and monetized benefits relevant to healthcare systems and funders. The ROI analysis focused on 171 program participants over 6 months, with baseline and follow-up survey data available (including how many times they had visited the emergency department or been hospitalized in the past six months). An additional 239 participants who enrolled in the program were excluded from the ROI analysis due to incomplete pre- and post-intervention data. While detailed comparative data are limited, available information suggests that participants excluded from the analytic sample were broadly similar in demographic composition, though they may have differed in engagement levels and survey completion, which could introduce potential selection bias. These results, combined with evidence-informed estimates of avoided complications and improved medication adherence, were used to calculate the intervention’s financial return in a highly-impacted, under-resourced population.

The DSMP consisted of a standardized six-week, peer-led education series facilitated by trained CHWs. Workshops were delivered in English, Spanish, and Korean, and were designed for adults living in underserved neighborhoods in Melrose, Morrisania, and East Harlem. These communities were selected based on elevated rates of diabetes and persistent barriers to care, including limited access to health services, high rates of poverty, and food insecurity. In addition to delivering the evidence-based curriculum, CHWs used a standardized screening tool for HRSNs to generate culturally appropriate referrals to community resources within the NowPow digital platform. The program was implemented by three partner organizations, with centralized training, quality assurance, and technical support from the Health Department to ensure consistent quality and delivery across sites.

Cost data for this analysis were derived from the time-driven, activity-based costing model developed during the pilot. Direct implementation costs were estimated at **$744 per participant**, encompassing CHW and administrative staffing, workshop facilitation, participant recruitment, HRSN screening and referrals, materials, and training across partner organizations. For the 171 participants who completed both a baseline and follow-up survey, the total direct program cost was **$127**,**224**. To better reflect the full investment required to implement and sustain the program, several additional operational costs were considered. These included the cost of CHW turnover and the administrative overhead for partner organizations to support recruitment, data collection, coordination, and reporting activities throughout the intervention.

CHW turnover was modeled at 30% annually, consistent with national estimates,^8^ and the cost to replace one CHW was conservatively estimated at 50% of a $40,000 annual salary, inclusive of employee benefits.^9,10^ For two full-time CHWs, the resulting yearly turnover cost was $12,000. Data collection, entry, and reporting responsibilities assigned to the participating CBOs were valued at $40,000 annually, based on average labor costs and administrative overhead reported in the community health literature. These indirect costs brought the total program investment to $179,224. The total program costs are summarized in Table 1.

**Table 1:** Total Program Costs for One City (Year 1, 171 Participants)

| Cost Category | Unit Cost / Rate | Quantity / Assumption | Total Cost (USD) |
| --- | --- | --- | --- |
| **Direct Implementation Costs** |  |  |  |
| Per-participant program delivery cost | \$744 per participant | 171 participants | \$127,224 |
| **Indirect and Operational Costs** |  |  |  |
| CHW turnover (30% of 2 FTE CHWs) | \$20,000 per turnover (50% of annual salary) | 2 CHWs × 30% = 0.6 FTE turnover | \$12,000 |
| Data collection and reporting (CBOs) | Fixed annual administrative estimate | Reporting, QA, staff work for 1 year | \$40,000 |
| **TOTAL PROGRAM COST (Fully Loaded)** | | | **\$179,224** |

To estimate benefits, four monetizable categories were modeled: avoided diabetes complications, avoided emergency department (ED) visits, avoided hospitalizations, and gains in quality-adjusted life years (QALYs) due to improved medication adherence. Avoided complications were valued at 20% of the annual average cost of treating diabetes-related conditions, estimated at $12,000 per person per year.^11^ This yielded a benefit of $2,400 per participant, or $196,800 across the 171 completers. Data from the Health Department’s 2019 evaluation indicated a reduction in self-reported ED visits from 149 to 82 in successive surveys comparing the six-month periods before and after the intervention among a cohort of 171 participants with complete data. This represented 67 avoided ED visits at a cost of $1,389 per visit, for a total savings of $93,063. Similarly, self-reported hospitalizations declined from 93 to 24 during the same period, yielding 69 avoided hospitalizations valued at $9,600 each, for a total of $662,400 in savings.

In addition to reductions in acute care utilization, the analysis also incorporated improvements in medication adherence and their associated impact on long-term health outcomes. Based on findings from recent cost-effectiveness studies, improvements in adherence attributable to DSMP participation generate an average QALY gain of 0.077 per person. Valued at the commonly accepted public health benchmark of $50,000 per QALY, this translates to $3,850 per participant, or $658,350 across the 171 participants. A summary of the monetized benefits is shown in Table 2.

**Table 2:** Monetized Benefits of NYC DSMP (Year 1, 171 Participants)

| **Benefit Category** | **Unit Value (USD)** | **Quantity / Assumption** | **Total Benefit (USD)** |
| --- | --- | --- | --- |
| **Avoided Diabetes Complications** | \$2,400 per participant | 20% reduction of \$12,000 avg. annual cost × 171 | \$410,400 |
| **Avoided Emergency Department Visits** | \$1,389 per visit (CDC estimate) | 67 avoided visits (scaled from OneCity data) | \$93,063 |
| **Avoided Hospitalizations** | \$9,600 per hospitalization (AHRQ estimate) | 69 avoided hospitalizations (scaled) | \$662,400 |
| **QALY Gains from Medication Adherence** | \$3,850 per participant | 0.077 QALY × \$50,000 × 171 participants | \$658,350 |
| **TOTAL MONETIZED BENEFIT** | | | **\$1,824,213** |

To account for model uncertainty, a univariate sensitivity analysis was conducted on key assumptions, including QALY valuation, hospitalization rates, and indirect cost estimates. Even under conservative scenarios with reduced hospitalization impact and QALY valuation, the program maintained a positive ROI, supporting its cost-effectiveness and potential scalability within public health systems serving highly impacted populations. Exhibit 1 shows the impact of the analysis on ROI estimates.

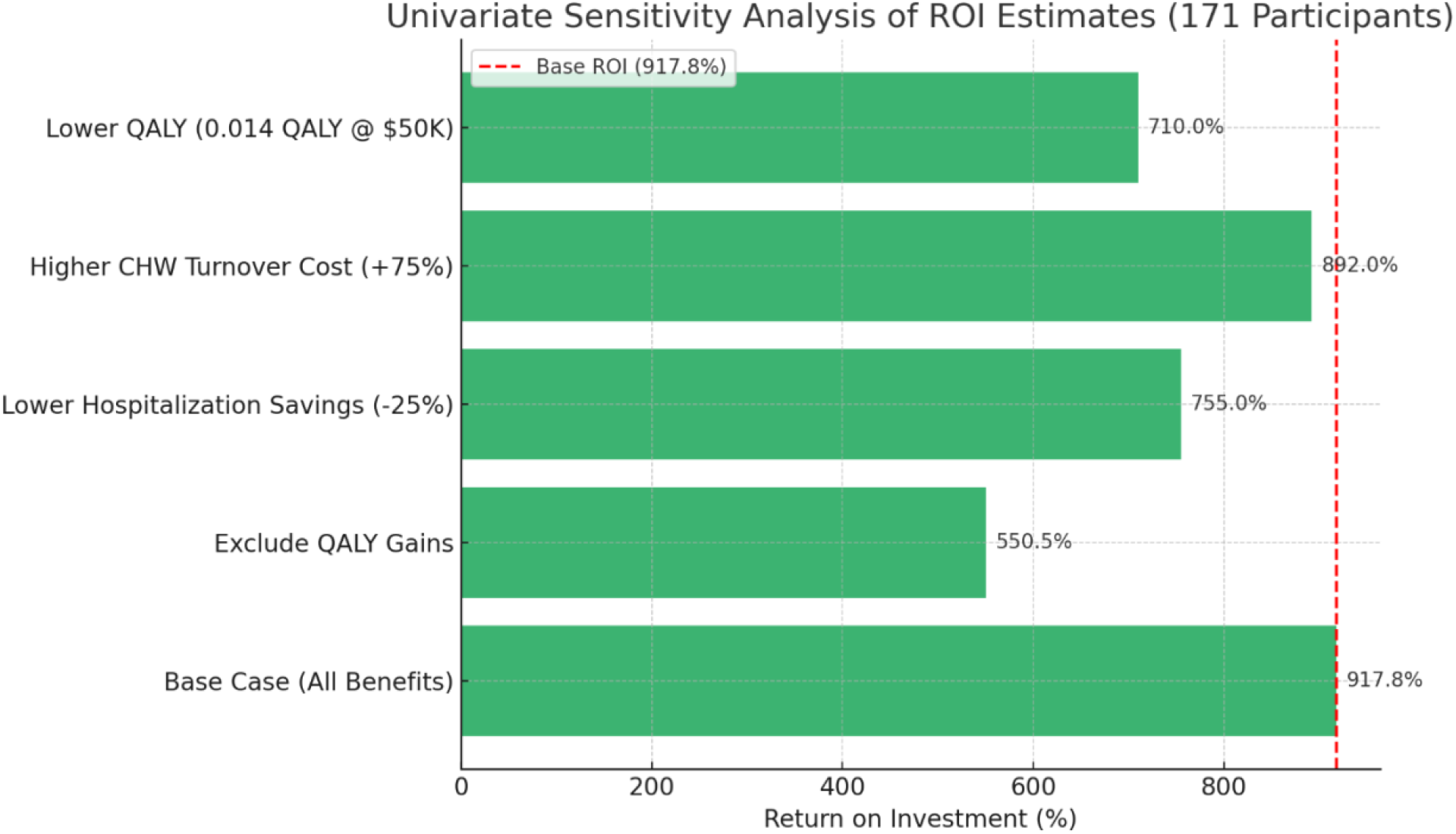

## Results

The ROI analysis found that the DSMP pilot program, enhanced with social needs screening and referrals, generated a substantial economic return within its first year of implementation. After accounting for all direct and indirect program costs, including workforce turnover and administrative reporting activities, the total cost of delivering the program to 171 participants was estimated at **$179**,**224**. Monetized benefits, derived from self-reported reductions in emergency department visits and hospitalizations, as well as modeled improvements in diabetes-related complications and medication adherence, resulted in a total estimated benefit of **$1**,**824**,**213**. These gains were based on participant-level impacts scaled from existing program data and supported by peer-reviewed literature.

The resulting **net benefit** was **$1**,**644**,**989**, yielding a **return on investment (ROI) of 918%**. This indicates that for every $1 invested in the DSMP, the program returned approximately $**10** in economic value to various stakeholders across society, especially insurers. When excluding the quality-adjusted life year (QALY) gains from improved medication adherence, the ROI remained strong at **551%**, underscoring the program’s cost-effectiveness even under conservative assumptions. The sensitivity analysis confirmed the robustness of these results. Even when modeling reductions in hospitalization impact, increases in workforce turnover costs, or lower QALY values, the program consistently delivered positive net returns. These findings suggest that the DSMP is both financially and clinically impactful, offering an efficient use of public health resources in highly impacted communities, specifically those with a very high ED and hospital admission rates at baseline. An ROI summary is shown in Table 3.

**Table 3:** ROI Summary Table.

| ROI Scenario | Total Benefits | Total Costs | Net Benefit | ROI (%) |
| --- | --- | --- | --- | --- |
| **Without QALY Gains** | \$1,165,863 | \$143,008 | \$986,639 | **550.5%** |
| **With QALY Gains** | \$1,824,213 | \$143,008 | \$1,644,989 | **917.8%** |

## Discussion

The results of this analysis demonstrate that DSMP, integrated with social needs screening and referral piloted by the Health Department and Health People Inc, The Mexican Coalition for the Empowerment of Youth and People, and the Korean Community Services of Metropolitan New York Inc., yielded a strong return on investment (ROI) during the one-year period of operation. The majority (over 80%) of the program participants reported housing instability. Using a fully loaded cost model that includes direct implementation expenses, workforce turnover, and administrative reporting activities, the program generated an ROI of over 500% excluding quality-adjusted life years (QALYs) and over 900% including QALYs attributable to improved medication adherence. These findings indicate that the program not only achieved its intended public health outcomes (e.g., increased self-efficacy, reduced utilization, and improved diabetes knowledge) but also yielded substantial economic value for public payers and the health system.

Importantly, the analysis was grounded in real-world implementation data from the pilot study, which documented improved self-reported health behaviors, reduced ED visits, and decreased hospital admissions among DSMP participants. These outcomes are consistent with prior research demonstrating the clinical and economic impact of community-based diabetes interventions, particularly those facilitated by CHWs^.12,13^ Nationally, it has been shown that a 1% reduction in HbA1c levels can lead to annual healthcare savings of $429 to $736 per person,^14^ and that CHW-facilitated chronic care models are associated with significant reductions in inpatient utilization, particularly in populations with low household incomes.^15,16^

This study contributes to the existing literature by offering a detailed ROI model that incorporates not only clinical cost savings but also structural and systems-level investments, such as health IT infrastructure and administrative overhead, that are often overlooked in short-term economic evaluations. The inclusion of QALY gains tied to improved medication adherence adds another vital dimension. While often excluded from early-stage evaluations, QALYs offer a standardized metric for assessing improvements in health-related quality of life. Even when modeled conservatively at 0.077 QALYs per participant, the additional value generated was significant, contributing over $300,000 in economic return. Importantly, the structure and findings of this model are relevant beyond urban settings; similar economic and health impacts may be observed in rural communities and other under-resourced regions that experience comparable chronic disease burdens, workforce shortages, and demographic disparities. In such contexts, where access barriers, limited specialty care, and social determinants of health play a substantial role, community-based, CHW-led interventions may offer a particularly scalable and cost-effective strategy for addressing persistent inequities in diabetes outcomes. The findings are additionally striking given that the population reported high levels of housing instability. Public health tools are essential in mitigating the vicious cycle between unstable housing and chronic disease managementl. Our findings suggest government-supported DSMP may mitigate the impacts of diabetes on people facing housing instability.

Beyond its financial impact, the DSMP pilot was closely aligned with public health priorities focused on advancing health equity, strengthening the community-based workforce, and improving connections between clinical services and neighborhood-level support systems. The program was strategically implemented among people, many with unstable housing, in neighborhoods with historic segregation, high diabetes prevalence and persistent structural barriers to care, including limited access to healthcare, language barriers, and socioeconomic disadvantage. Participants predominantly represented priority populations, including Black and Latino adults, older adults, and English language learners. Further, the largest payer in this group was Medicaid (72%) followed by uninsured (16)%. By engaging CHWs from the communities being served, the program fostered cultural relevance and trust while simultaneously investing in the development of a diverse public health workforce equipped to support chronic disease management at the community level.

These findings also carry implications for Medicaid policy and managed care organizations, particularly in the context of value-based payment models. The financial return demonstrated in this study supports the case for incorporating CHW-led diabetes self-management programs enhanced by social needs screening and referral into managed care benefit design, capitated payment structures, or Section 1115 demonstration waivers. Our results make a strong case for including DSMP as a covered benefit by both commercial health plans, such as those covering city workers, and Medicaid managed care. Additionally, given the results in a population facing housing instability, DSMP can be integrated into housing access and affordability initiatives.

Additionally, the economic case for policy makers to support the sustainable of such programs becomes even more compelling when broader social benefits, such as increased workforce productivity, reduced caregiver burden, and improved patient satisfaction, are considered. However, these were not included in this ROI model.

Despite this analysis’s strengths, there are significant limitations that merit consideration. First, the evaluation relied in part on self-reported data for ED visits, hospitalizations, and health behaviors, which may be subject to recall or reporting bias. Second, the absence of a control group limits causal attribution of outcomes to the DSMP intervention alone. Third, some cost and benefit parameters, particularly QALY valuation and hospitalization costs, were derived from the national literature rather than NYC-specific claims data, potentially introducing uncertainty into the model. However, sensitivity analyses demonstrated that the ROI remained robust under a range of conservative assumptions.

Finally, the analysis reflects only one year of program implementation in three CBOs. While the short-term ROI is impressive, the full value of diabetes self-management support may become even more apparent over the long term as participants maintain behavior changes, avoid long-term complications, and remain engaged in care. Future work should focus on multi-year cost trajectories, claims-based validation of utilization reductions, and the inclusion of broader social return-on-investment (SROI) metrics that account for social determinants of health and quality of life.

## Conclusion

As cities, counties, and states explore scalable interventions to address chronic disease and health inequities in areas facing housing instability and other social needs, DSMP offers a strong return on investment. DSMP can be integrated with other evidence-informed public health interventions, such as telephonic self-management.^17^ Central to DSMP’s effectiveness are peer leaders and community health workers (CHWs), who are uniquely positioned to deliver high-quality, culturally responsive care that is both clinically impactful and cost-effective. By leveraging trusted community relationships, these essential workers extend the reach of traditional healthcare systems while reducing reliance on high-cost acute services. Future research should be conducted during the scale process as both local governments and payers invest in DSMP. Such studies at scale can assess DSMP’s long-term impact, validate costs using claims data, and quantify broader societal benefits, such as improved productivity, caregiver support, and community resilience.

## Data Availability

All data produced in the present study are available upon reasonable request to the authors

